# Multimodal semantic knowledge of emotion concepts in frontotemporal dementia

**DOI:** 10.64898/2026.01.12.26343893

**Authors:** Maxime Montembeault, Valentina Borghesani, Zachary A. Miller, Maria Luisa Mandelli, Janhavi Pillai, Sheila Tran, Rian Bogley, Carly Millanski, Zoe Ezzes, Buddhika Ratnasiri, Eleanor R. Palser, Hulya Ulugut, Kyan Younes, Michelle Shiota, Dacher Keltner, Alan Cowen, Maya L. Henry, Katherine P. Rankin, Virginia E. Sturm, Maria Luisa Gorno-Tempini

## Abstract

**Background:** Recent work has delineated the semantic behavioral variant of frontotemporal dementia (sbvFTD; or right temporal variant of FTD, which is thought to preferentially impair semantic knowledge for emotional concepts. However, this proposed core feature has not yet been empirically validated, and no clinical tool exists to assess it. Establishing reliable markers is essential to clinically differentiate sbvFTD from behavioral variant FTD (bvFTD), which is critical given their overlapping behavioral symptoms but divergent underlying pathologies. Furthermore, contrasting sbvFTD with semantic variant primary progressive aphasia (svPPA) can advance our understanding of semantic memory, revealing how the right and left anterior temporal lobes (ATLs) support emotion- versus tool-related knowledge, highlighting the graded, lateralized organization of the semantic system.

**Methods:** We studied 15 patients with sbvFTD, 15 with svPPA, 18 with bvFTD, and 37 healthy controls. A novel multimodal semantic battery, the Fear and Spider Test (FST), which assesses tool- and emotion-related concepts across word-based semantic associations, picture-based semantic associations, and sound-to-picture matching, was administered. Stimuli were matched on psycholinguistic and perceptual features, and emotional items were drawn from multicultural facial expressions validated with the Facial Action Coding System. Neural correlates of semantic performance were investigated using voxel-based morphometry.

**Results:** As expected, patients with both sbvFTD and svPPA showed greater deficits in all semantic tasks compared to controls and bvFTD, and bilateral anterior temporal lobe (ATL) volumes were broadly associated with performance across all semantic tasks. Interactions between modality and categories were necessary for the emergence of differences between sbvFTD and svPPA and right and left ATL atrophy: performance on the Words–Tools condition was more impaired in svPPA and correlated with left ATL volume, while performance on the Pictures–Emotions condition was more impaired in sbvFTD and correlated with right ATL volume.

**Conclusion:** The FST provides the first clear dissociation of sbvFTD from bvFTD, a distinction of critical clinical importance given their divergent pathological substrates and the absence of frontotemporal lobar degeneration specific biomarkers. The study also refines our understanding of semantic memory: contrasting sbvFTD with svPPA reveals complementary roles of the right and left ATLs in supporting emotion and tool knowledge, underscoring the graded, lateralized organization of the semantic system.

## 1. Introduction

Patients with progressive degeneration of the right anterior temporal lobe (ATL) have recently been the focus of proposed diagnostic criteria under the term “semantic behavioral variant frontotemporal dementia” (sbvFTD) (Younes et al., 2022). The core clinical symptom profile described loss of empathy, impaired recognition and naming of familiar individuals, and rigid or compulsive behaviors. In the same line, a dedicated international working group on this topic has proposed that the three primary domains of impairment in this clinical syndrome were related to (i) multimodal knowledge of non-verbal information including people, living beings, landmarks, flavors/odors, sounds, bodily sensations, emotions and social cues; (ii) socio-emotional behavior encompassing emotion expression, social response and motivation; and (iii) prioritization for focus on specific interests, hedonic valuation and personal preferences (Ulugut, Bertoux, et al., 2024; Ulugut, Younes, et al., 2024a). In the past, these cases have also been labeled as right semantic dementia, right semantic variant primary progressive aphasia (svPPA), or right temporal variant FTD, or simply diagnosed as behavioral variant FTD (bvFTD) or svPPA (Ulugut, Bertoux, et al., 2024; Ulugut, Younes, et al., 2024b; Younes et al., 2022). BvFTD, on the other hand, is characterized by early changes in personality, social behavior, and executive functioning, such as disinhibition, apathy, loss of empathy, and stereotyped behaviors, typically associated with frontoinsular atrophy (Rascovsky et al., 2011). Finally, svPPA is marked by progressive degradation of verbal semantic knowledge, particularly impaired single-word comprehension and confrontation naming, usually associated with left-lateralized ATL atrophy (Gorno-Tempini et al., 2011).

Although sbvFTD appears to fall between svPPA and bvFTD, sharing semantic and linguistic symptoms with the former, and socioemotional and behavioral symptoms with the latter, the diagnostic criteria for neither of these clinical syndromes captures the clinical changes in sbvFTD. A recent study showed that less than 30% of right ATL-predominant patients met formal diagnostic criteria for either svPPA or bvFTD within the first three years of disease progression (Younes et al., 2022). Nonetheless, differentiating sbvFTD from bvFTD is essential due to its distinct probability of underlying pathology: while sbvFTD shares Frontotemporal Lobar Degeneration (FTLD)-TDP type C pathology with svPPA in most cases (∼84%), bvFTD is associated with more heterogeneous pathologies, including FTLD-TDP types A/B and FTLD-tau (Perry et al., 2017; Spinelli et al., 2017). Accurate diagnostic classification thus has critical implications for clinical management and the development of targeted therapies. Despite this clinical relevance, very few studies have prospectively compared sbvFTD samples with bvFTD and svPPA, and reliable tools for diagnosing sbvFTD in clinics and research studies are lacking.

As proposed by Younes et al. (2022), a core feature of sbvFTD may be the degradation of nonverbal and socio-emotionally relevant conceptual knowledge. A recent meta-analysis in healthy populations has documented the link between emotion recognition and empathy deficits (Qiao et al., 2025), and a similar mechanism might be at play in sbvFTD. Specifically, patients with sbvFTD might no longer reliably associate nonverbal social cues, such as facial expressions, body language, or tone of voice, with their intended emotional meaning, which may, in turn, help explain why their behaviors can sometimes appear insensitive, inappropriate, or socially out of place. The core importance of semantic deficits in sbvFTD is not surprising when considering neuroscientific accounts of semantic memory. Semantic memory appears to be supported by ATL regions, bilaterally, with graded variations in function reflecting differential patterns of connectivity (M. A. Lambon Ralph et al., 2001, 2010; M. A. L. Lambon Ralph et al., 2017; Rice et al., 2018). In terms of sensory modality, whereas the left ATL binds verbal features into semantic knowledge through strong connections with left-sided linguistic networks, the right ATL may be more involved in representing non-verbal semantic knowledge through its prominent connections with right-sided visual networks. This is consistent with previous clinical studies showing that svPPA patients with predominant left ATL atrophy are significantly more impaired in verbal semantic tasks (Binney et al., 2016; Chen et al., 2018; Snowden et al., 2018; Woollams & Patterson, 2018), while patients with predominant right ATL atrophy are significantly more impaired in visual semantic tasks (Luzzi et al., 2017; Woollams & Patterson, 2018).

Beyond the sensory modality effect, the semantic category of concepts also influences both the laterality of brain correlates and the degree of semantic impairment. In neurodegenerative diseases, semantic processing of living stimuli has been linked to gray matter volume in the right ATL, whereas the processing of non-living items such as tools and household objects has been associated with volume in the left posterior middle temporal gyrus (Brambati et al., 2006). More specifically, action- or motor-based knowledge of tool use has been shown to rely on a left-lateralized network, including the inferior and superior parietal lobules and premotor regions. This conclusion is supported by many studies, including an activation likelihood estimation meta-analysis of 70 neuroimaging contrasts comparing manipulable versus non-manipulable tools (Borghesani et al., 2019; Canessa et al., 2008; Ishibashi et al., 2016; Wurm & Lingnau, 2015). However, emotion concepts remain underexplored within this framework. It is hypothesized that, through its connections with predominantly right-hemisphere emotional networks and given that emotional information is often processed through non-verbal inputs (e.g., facial expressions, affective prosody, bodily movements), the right ATL serves as a core hub for emotional semantic knowledge. This hypothesis is supported by clinical studies showing worse emotion recognition in patients with right ATL degeneration compared to those with svPPA (Irish et al., 2013; Rosen et al., 2004), although these studies did not use semantic tests per se. It is also supported by studies of emotion perception suggesting a relative lateralization to the right hemisphere, including regions of the temporal lobe (Schirmer & Adolphs, 2017).

While research into the neurobiology of semantic memory has enhanced our understanding of modality (e.g., verbal vs. non-verbal) and category (e.g., tools vs. emotions) effects, these advancements have not yet been integrated into clinical practice for patients with sbvFTD. As a result, they tend to overlook other sensory modalities, such as auditory input, and neglect semantically relevant categories like emotional concepts. Emotion recognition tests, while targeting affective content, are typically limited to matching basic facial expressions to labels and therefore do not evaluate the conceptual understanding of emotions. Furthermore, they are rarely multimodal, and do not encompass a wide range of emotions, including recently validated positive emotions such as pride or awe (Cordaro et al., 2020; Cowen et al., 2019). Finally, empathy questionnaires are mostly informant-rated and assess real-life behavior rather than semantic knowledge as assessed in semantic association neuropsychological tasks.

To address these limitations, we developed the Fear & Spider Test (FST), a novel multimodal semantic battery contrasting emotion and tool concepts across three modalities: sound-to-picture, words, and pictures. The battery was carefully designed to include multicultural facial expressions validated with the Facial Action Coding System (FACS), covering both positive and negative emotions (Cordaro et al., 2020; Cowen et al., 2019). Subtests were rigorously matched on multiple psycholinguistic variables. In this first prospective study of sbvFTD since the publication of its proposed diagnostic criteria, we examined 15 sbvFTD patients, 15 patients with svPPA, 18 patients with bvFTD, and 37 healthy controls (HC). We aimed to determine the effects of modality and semantic category on performance in the FST across clinical groups. We hypothesized that: (1) the battery would successfully differentiate the three patient groups based on their distinct semantic profiles; (2) sbvFTD patients would exhibit widespread semantic impairments, particularly in nonverbal modalities and for emotion-related concepts (3) svPPA patients would show extensive semantic deficits, especially for the verbal modality and tool-related concepts; and (4) bvFTD patients would be relatively preserved across tasks, except in nonverbal subtasks involving emotional concepts given the recognized emotion recognition deficits in this population (Bertoux et al., 2015). A second goal of our study was to investigate the brain regions associated with semantic performance in different modalities and semantic categories of the Fear & Spider Test. We hypothesized that performance across all subtasks would correlate with the ATLs, with a graded lateralization pattern: left-predominant involvement for verbal and tool-related tasks, and right-predominant involvement for nonverbal and emotion-related tasks. We also expected an interaction between impairment in verbal semantic knowledge of tools and visual semantic knowledge of emotions in svPPA and sbvFTD, respectively, and their association with lateralized ATL atrophy.

## 2. Methods

### 2.1. Participants

Patients were recruited through the University of California, San Francisco (UCSF) Edward and Pearl Fein Memory and Aging Center (MAC) between 2019 and 2023. The diagnosis was made after a comprehensive evaluation (neurologic history and examination, standardized neuropsychological and language evaluations) and a review of this evaluation at a consensus diagnostic meeting at the UCSF MAC. Patients with bvFTD (n=18) fulfilled the Rascovsky consensus diagnostic criteria (Rascovsky et al., 2011). Patients with svPPA (n=15) fulfilled the Gorno-Tempini consensus diagnostic criteria (Gorno-Tempini et al., 2011). Finally, patients with sbvFTD (n=15) fulfilled the Younes proposed diagnostic criteria (Younes et al., 2022). HCs (n=37) who were neurologically normal as attested by a neurologic examination, neuropsychological evaluation, and MRI were also included in the sample as a comparison group. All participants gave written consent, and the study was approved by the institutional review board.

### 2.2. Procedure

#### 2.2.1. General cognitive and language assessment

All participants underwent a broad neuropsychological battery and speech and language tests, as previously described (Gorno-Tempini et al., 2004; Kramer et al., 2003). To assess disease severity, we used the FTLD clinical dementia rating scale (FTLD-CDR) total and sum of boxes scores (Knopman et al., 2008).

#### 2.2.2. Fear & Spider test (FST): A new multimodal semantic battery to assess knowledge of emotions and tools

The battery evaluates the semantic knowledge of 28 concepts across two semantic categories (14 emotions and 14 tools) that can be presented in three modalities (word, picture, or sound). It includes three subtasks, each containing 28 items (Figure 1).

**Figure 1.**
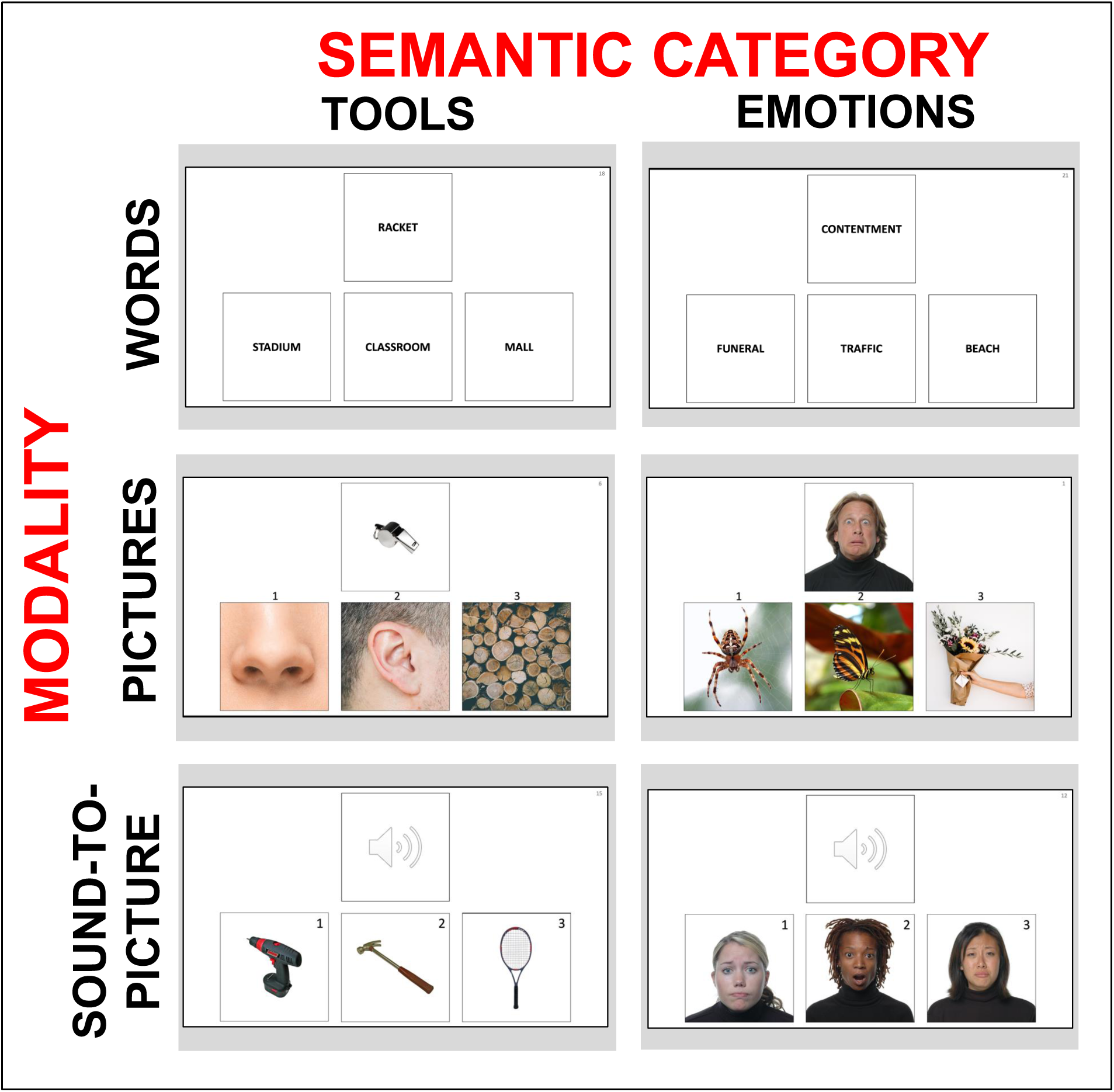
Structure of the Fear & Spider Test (FST). One example stimulus is presented for each modality & semantic category combination.

##### 2.2.2.1. Stimuli selection

The picture stimuli for tools were chosen from the Bank of Standardized Stimuli, a resource offering a comprehensive collection of well-characterized, high-quality photographs featuring various objects against a white background (Brodeur et al., 2010). The picture stimuli for emotions were chosen from a study by Cordaro and colleagues, encompassing 18 facial-bodily expressions by diverse actors on a white background (Cordaro et al., 2020). This specific set was selected due to its inclusion of more positive emotions than typical sets (such as amusement, contentment, sympathy), elicited through a detailed procedure. More precisely, two researchers certified in the Facial Action Coding System guided eight paid posers, all U.S. citizens, with muscle-by-muscle instructions to configure the states based on anatomical movements documented in independent research. Four additional picture stimuli for the emotions of awe, love, enthusiasm, and pride were obtained through unpublished work by author MS and her team. This allowed us to select 7 negative and 7 positive emotions in our set of emotions. Sound stimuli for tools were selected from various sources on the internet. Finally, sound stimuli for emotions were selected from a study by Cowen and colleagues, encompassing well-characterized emotion stimuli conveyed by brief human vocalizations or vocal bursts across 24 different emotions (Cowen et al., 2019).

##### 2.2.2.2. Task development

After selecting stimuli (words, pictures, and sounds) for potential tool and emotion concepts. We extracted the psycholinguistic characteristics of the words (length, lexical frequency, and age of acquisition) from English databases. For the pictures and sounds, we conducted a pilot survey with 15 participants to obtain familiarity ratings. Based on these data, we selected stimuli and created two lists, each containing 14 tool-related and 14 emotion-related concepts, matched on all these variables. This was done to ensure that statistical analyses investigating the effect of semantic category within this study would be controlled for all psycholinguistic characteristics of the stimuli.

In preparation for the development of the Semantic Associations – Words and Pictures tasks, we also generated one concept associated with each primary tool or emotion concept, along with two distractor concepts that were not associated with the primary concept. We extracted the same psycholinguistic variables and collected pilot survey ratings for these associated and distractor concepts as we did for the main concepts. Similarly, as with the main concepts, we then ensured that the lists of targets (concepts associated with the primary tool or emotion concepts) and distractors (unassociated concepts) were matched between the tool and emotion categories.

Once all items were created, a final pilot survey was conducted with 42 participants to confirm that all items were adequate, successfully completed, and well understood by healthy older adults at a minimum success rate of 70%. As all items met this criterion, they were retained for further use in the Fear & Spider Test.

##### 2.2.2.3. Administration of the subtasks

In the sound-to-picture matching subtask, participants had to listen to a sound and identify which picture, among three options, matched the sound. It was specified to the participants that they needed to match the sound and picture based on the expressed emotion rather than on the visual and auditory characteristics of the individuals depicted in the stimuli. Participants had the option to request an additional playback.

In the Semantic associations - Words subtask, participants were required to examine a word (representing one of the 28 concepts) and choose, from three options, the word of a context that would be the most associated with the target concept (for example, the word shovel with the word blizzard). Words were read aloud by the examiner to accommodate potential reading impairments.

In the Semantic associations - Pictures subtask, participants were required to make the exact same associations as in the Semantic associations – Words subtask but using pictures stimuli instead of words.

In each subtask, stimuli alternated between one emotion and one tool, enabling the investigation of the semantic category’s effect. The test battery included two counterbalanced presentation orders to allow for the exploration of modality effects (counterbalanced sequences of the Semantic associations – Words and Pictures subtasks (14 stimuli from subtask A, 28 items from subtask B, 14 stimuli from subtask A). Before each subtask, two practice items were provided and explained if necessary to ensure participants’ full comprehension of the task. There was no time limit set for each item. All subtasks provide a total score (/28), an emotion concepts score (/14) and a tool concepts score (/14).

#### 2.2.3. Neuroimaging protocol

Participants were scanned with a Siemens 3-Tesla Prisma scanner using a body transmit coil and a 64-channel receive head coil. The structural MRI included a T1-weighted 3D magnetization prepared rapid acquisition gradient echo (MPRAGE) acquired with 160 sagittal slices, TE/TR/TI = 2.9/2300/900 ms, flip angle = 9°, isotropic voxel with size of 1 mm, field-of-view = 256 × 256 mm^2^, matrix = 256 × 256. Structural MRI data were preprocessed using the Computational Anatomy Toolbox (CAT12; dbm.neuro.uni-jena.de/cat) running under MATLAB. CAT12 classifies T1-weighted data as gray matter (GM), white matter, or cerebrospinal fluid using an improved segmentation approach compared to the traditional unified segmentation (Ashburner & Friston, 2005), based on an adaptive maximum a posteriori. GM probability maps were then nonlinearly normalized to the Montreal Neurologic Institute (MNI) space using Diffeomorphic Anatomical Registration Through Exponentiated Lie Algebra (DARTEL)(Ashburner, 2007), and modulated by the Jacobian determinant of the deformations derived from the spatial normalization. Modulated GM images were smoothed with an isotropic Gaussian kernel of 8 mm full width at half maximum (FWHM).

### 2.3. Statistical analyses

First, to assess group differences in performance across modalities, semantic categories, and experimental conditions in the Fear & Spider Test, we conducted three separate mixed-design ANCOVAs with group (HC, svPPA, sbvFTD, bvFTD) as the between-subjects factor. The within-subject factors were: (1) modality (Words, Pictures, Sound-to-picture), (2) semantic category (Emotions, Tools), and (3) condition (six task combinations crossing modality and category). All models controlled for age, sex, and education, and for comparisons between patient groups, FTLD-CDR Total score was additionally included as a covariate. Significant interactions were followed by simple main effects analyses and Bonferroni-corrected post hoc comparisons. Effect sizes were reported using partial eta squared (η²).

Second, to determine the GM correlates the FST subtasks, we used a voxel-based morphometry (VBM) approach to examine the correlation between each voxel of the whole brain and FST subtasks. We entered each subtask in a regression model as a variable of interest, with the normalized, modulated, smoothed GM images as inputs and including age, sex, FTLD CDR Total scores, and total intracranial volume as covariates of no interest. Contrasts were set to examine the hypothesis that reduced semantic performance would be associated with decreased GM volume. These association analyses were conducted on all participants.

Third, to investigate our a priori hypothesis of an interaction between svPPA and sbvFTD patients based on modality and semantic category, we conducted a series of analyses focusing on performance in the Words–Tools and Pictures–Emotions conditions. Although the primary mixed-design ANCOVAs described above assessed group differences across all modalities, categories, and conditions, their complexity and the small size of individual patient groups limited statistical power to detect targeted cross-over effects. Therefore, this aim was addressed using a planned, hypothesis-driven analytical approach. We first ran a 2 (Group: svPPA, sbvFTD) × 2 (Condition: Words–Tools, Pictures–Emotions) mixed-design ANCOVA, controlling for sex, to examine group-by-condition interaction effects. Post hoc comparisons were used to assess within- and between-group differences. To examine brain–behavior relationships beyond categorical diagnoses, we performed two multiple linear regression analyses across the full semantic dementia (SD) spectrum, using Words–Tools and Pictures–Emotions performance as dependent variables, and left and right ATL gray matter volumes as predictors, while controlling for age, sex, and total intracranial volume. We then computed a semantic index reflecting relative performance on the two tasks (higher values indicating greater impairment in Words–Tools), and compared groups on this index using ANCOVA, controlling for sex. Finally, we calculated a laterality index of ATL atrophy (positive values indicating left-dominant atrophy) and assessed its relationship with the semantic index using a partial correlation, adjusting for age, sex, and total intracranial volume.

## 3. Results

### 3.1 Description of participants (Table 1; Figure 2)

The demographic and clinical presentation summary of HC, sbvFTD, svPPA, and bvFTD patients is shown in Table 1. There was significant differences in sex distribution among the groups, with the HC and sbvFTD groups being more predominantly female and the svPPA and bvFTD groups being more predominantly male. There were also statistically significant age differences, with HC patients being older than those in the svPPA and bvFTD groups. Education levels were equivalent across groups. Regarding disease severity, svPPA patients had significantly lower FTLD CDR Total scores compared to bvFTD, but all groups presented equivalent FTLD CDR Box Scores. All analyses were controlled for sex, age, education and FTLD CDR Total scores (only between patients’ contrasts for FTLD CDR Total scores). The pattern of atrophy in each patient group is presented in Figure 2.

**Figure 2.**
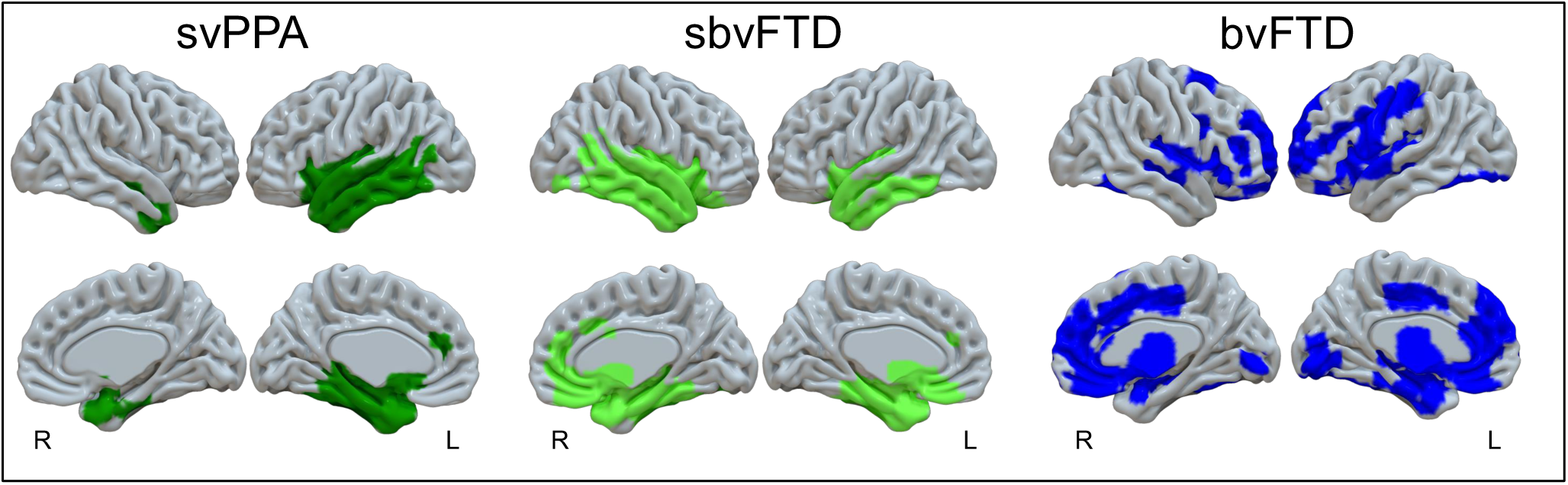
Gray matter atrophy patterns in each clinical group (p < .05 FWE corrected voxel-level for svPPA and sbvFTD and p < .05 FWE corrected cluster-level for bvFTD)

**Table 1.** Demographics, disease severity, language, and cognitive data.

|  | **HC**<br>**n=37** | **svPPA**<br>**n=15** | **sbvFTD**<br>**n=15** | **bvFTD**<br>**n=18** | **p** |
| --- | --- | --- | --- | --- | --- |
| **Demographics** |  |  |  |  |  |
| Sex (F/M) | 21/16 | 3/12<sup>a</sup> | 11/4<sup>b</sup> | 4/14<sup>a,c</sup> | <.01* |
| Age | 76.8 (7.0) | 68.3 (7.9)<sup>a</sup> | 71.6 (8.3) | 68.6 (7.6)<sup>a</sup> | <.001* |
| Years of education | 17.1 (2.1) | 16.8 (2.0) | 15.6 (2.5) | 17.3 (2.1) | .107 |
| **Disease severity** |  |  |  |  |  |
| FTLD CDR Total | 0.0 (0.0) | 1.1 (0.5)<sup>a</sup> | 1.3 (0.6)<sup>a</sup> | 1.5 (0.5)<sup>a,b</sup> | <.001* |
| FTLD CDR Box score | 0.0 (0.0) | 6.3 (2.9)<sup>a</sup> | 7.8 (4.0)<sup>a</sup> | 8.3 (2.9)<sup>a</sup> | <.001* |
| **Neuropsychological data** |  |  |  |  |  |
| Stroop interference (Number correct) | 52.2 (17.0) | 41.1 (15.9) | 38.6 (17.8) | 26.8 (16.7)<sup>a</sup> | <.001* |
| Modified Trail Test (s) | 30.1 (14.0) | 44.8 (33.1) | 61.3 (37.8)<sup>a</sup> | 65.0 (36.2)<sup>a</sup> | <.001* |
| Benson Figure Copy (/17) | 15.5 (0.8) | 15.4 (0.9) | 14.9 (2.5) | 14.2 (3.0) | .143 |
| Benson Figure Recall (/17) | 12.5 (2.5) | 7.8 (5.4)<sup>a</sup> | 4.9 (6.1)<sup>a</sup> | 8.6 (4.4)<sup>a</sup> | <.001* |
| VOSP Number Location (/10) | 9.1 (1.1) | 8.4 (2.5) | 8.5 (2.1) | 8.3 (1.7) | .330 |
| Boston Naming Test (/15) | 14.6 (1.2) | 3.3 (3.2)<sup>a</sup> | 6.0 (3.7)<sup>a</sup> | 13.2 (1.9)<sup>b,c</sup> | <.001* |
| PPVT (/16) | - | 6.1 (3.7) | 10.0 (2.9)<sup>b</sup> | 14.5 (2.0)<sup>b,c</sup> | <.001* |
| Irregular word reading (/6) | - | 3.2 (1.5) | 5.2 (1.0)<sup>b</sup> | 5.7 (0.5)<sup>b</sup> | <.001* |
| CATS Affect Matching (/16) | - | 10.3 (2.6) | 9.6 (3.8) | 11.1 (3.2) | .512 |

a = significantly different from HC
b = significantly different from svPPA
c = significantly different from sbvFTD
CATS = Comprehensive Affect Testing System; F = female; FTLD CDR = Frontotemporal Lobar Degeneration Clinical Dementia Rating scale; M = male; PPVT; Peabody Picture Vocabulary Test; VOSP = Visual Object and Space Perception Battery

### 3.2 Behavioral results from the Fear & Spider Test (Table 2; Figure 3; Supplementary Figure 1)

#### 3.2.1. Group * Modality (Words vs Pictures vs Sound-to-picture)

There was a statistically significant interaction between group and performance depending on the modality (*F*(6, 152) = 5.367, *p* < .001, partial η2 = .175), controlling for age, sex and education.

**Figure 3.**
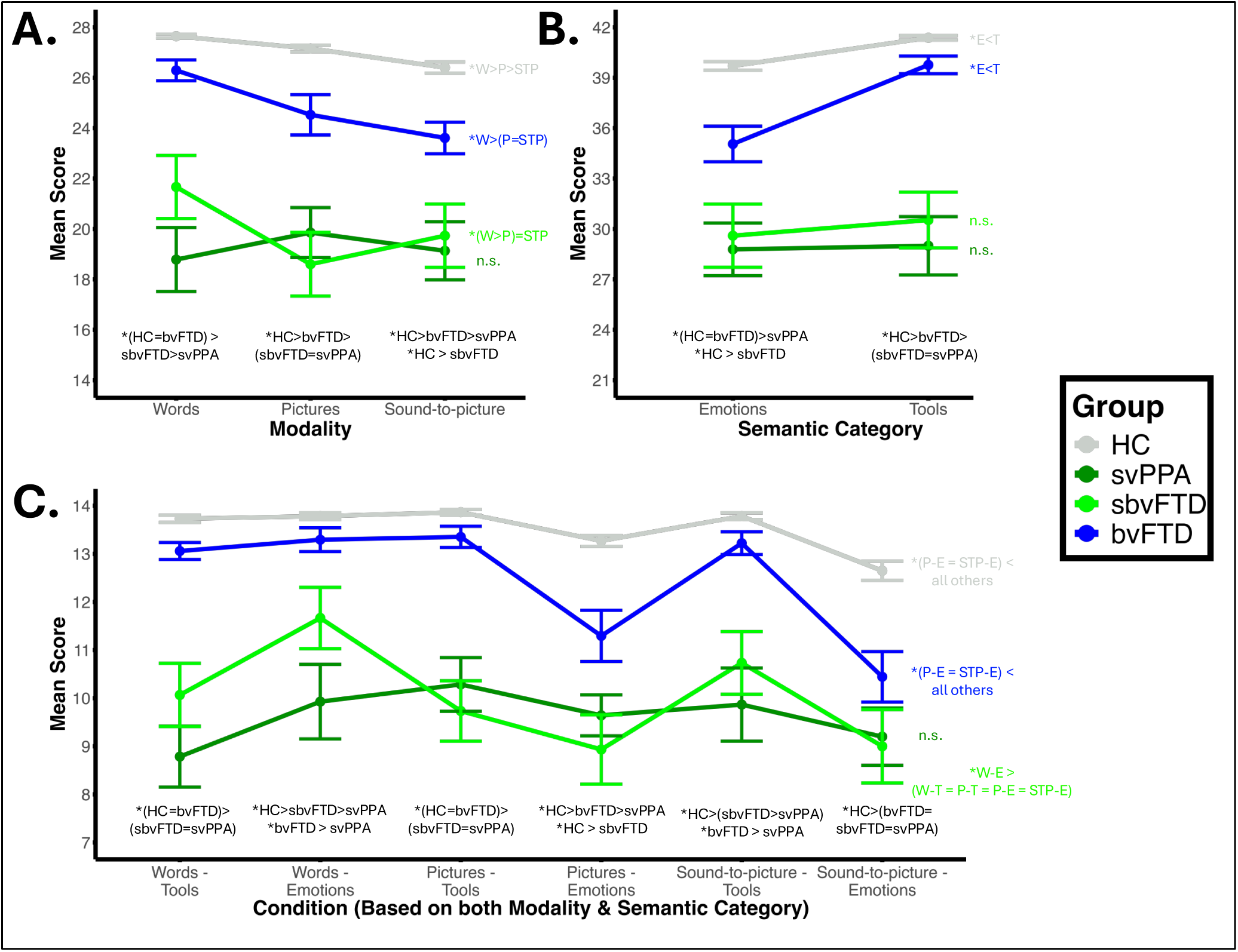
Behavioral Results from the Fear & Spider Test (FST): A) Interaction effect between group and modality; 2) Interaction effect between group and semantic category; 3) Interaction effect between group and condition.

**Table 2.** Behavioral results from the Fear & Spider Test (FST). Between-group differences are identified with letters next to the mean/standard deviations for each test conditions. Within-group differences are identified in the last row of each section.

|  | **HC**<br>**n=37** | **svPPA**<br>**n=15** | **sbvFTD**<br>**n=15** | **bvFTD**<br>**n=18** |
| --- | --- | --- | --- | --- |
| **Words (W; /28)** | 27.7 ± 0.5 | 18.8 ± 4.9 <sup>a</sup> | 21.7 ± 4.9 <sup>a,b</sup> | 26.3 ± 1.8 <sup>b,c</sup> |
| **Pictures (P; /28)** | 27.2 ± 0.8 | 19.9 ± 3.8 <sup>a</sup> | 18.6 ± 4.9 <sup>a</sup> | 24.5 ± 3.4 <sup>a,b,c</sup> |
| **Sound-to-picture (STP; /28)** | 26.4 ± 1.3 | 19.2 ± 4.6 <sup>a</sup> | 19.7 ± 4.9 <sup>a</sup> | 23.8 ± 2.6 <sup>a,b</sup> |
| **Within-group / Modality effects** | W>P>STP | n.s. | W>P | W>P;<br>W>STP |
| **Emotions (E; /42)** | 39.7 ± 1.6 | 28.8 ± 6.1 <sup>a</sup> | 29.6 ± 7.3 <sup>a</sup> | 35.1 ± 4.5 <sup>a,b</sup> |
| **Tools (T; /42)** | 41.4 ± 0.8 | 29.0 ± 6.7 <sup>a</sup> | 30.5 ± 6.4 <sup>a</sup> | 39.8 ± 2.2 <sup>b,c</sup> |
| **Within-group / Semantic category effects** | E<T | n.s. | n.s. | E<T |
| **Sound-to-picture – Emotions (STP-E; /14)** | 12.7 ± 1.2 | 9.2 ± 2.4 <sup>a</sup> | 9.0 ± 3.0 <sup>a</sup> | 10.5 ± 2.3 <sup>a</sup> |
| **Sound-to-picture – Tools (STP-T; /14)** | 13.8 ± 0.4 | 9.9 ± 3.1 <sup>a</sup> | 10.7 ± 2.5 <sup>a,b</sup> | 13.4 ± 0.9 <sup>b,c</sup> |
| **Pictures – Emotions (P-E; /14)** | 13.2 ± 0.7 | 9.6 ± 1.6 <sup>a</sup> | 8.9 ± 2.8 <sup>a</sup> | 11.3 ± 2.3 <sup>a,b,c</sup> |
| **Pictures – Tools (P-T; /14)** | 13.9 ± 0.4 | 10.3 ± 2.2 <sup>a</sup> | 9.7 ± 2.4 <sup>a</sup> | 13.4 ± 0.9 <sup>b,c</sup> |
| **Words – Emotions (W-E; /14)** | 13.8 ± 0.4 | 9.9 ± 3.0 <sup>a</sup> | 11.7 ± 2.5 <sup>a,b</sup> | 13.3 ± 1.1 <sup>b</sup> |
| **Words – Tools (WT; /14)** | 13.7 ± 0.5 | 8.8 ± 2.5 <sup>a</sup> | 10.1 ± 2.5 <sup>a</sup> | 13.1 ± 0.8 <sup>b,c</sup> |
| **Within-group / Condition effects** | (P-E = STP-E) <<br>all others | n.s. | W-E ><br>(STP-E, P-E, P-T, W-T) | (P-E = STP-E) <<br>all others |

a = significantly different from HC
b = significantly different from svPPA
c = significantly different from sbvFTD
n.s. = non-significant
W = words; P = pictures; STP = sound-to-picture
E = emotions; T = tools

In terms of simple main effects of groups, for the words modality, svPPA and sbvFTD performed significantly lower than HC (p < .001 for both), but bvFTD did not significantly differ from HC (p = .698). SvPPA and sbvFTD also performed significantly lower than bvFTD (p < .001 and p = .019, respectively). Finally, svPPA also performed significantly lower than sbvFTD (p = .008). For the pictures modality, all patients’ groups performed significantly lower than HC (p < .001 for svPPA and sbvFTD, p = 0.045 for bvFTD). SvPPA and sbvFTD also performed significantly lower than bvFTD (p < .001 for both), but svPPA and sbvFTD did not significantly differ (p = .999). For the sound-to-picture modality, all patient groups performed significantly lower than HC (p < .001 for svPPA and sbvFTD, p = .020 for bvFTD). SvPPA also performed significantly lower than bvFTD (p < .001), but sbvFTD and bvFTD (p = .057) and svPPA and sbvFTD (p = .222) did not significantly differ. Comparisons between the patient groups were additionally controlled for FTLD CDR Total scores.

In terms of simple main effects of modality, in HC, there was a statistically significant effect of modality on performance (*F*(2, 72) = 18.58, *p* < .001, partial η2 = .340). Post-hoc tests showed that performance in the words modality was significantly higher than in the pictures modality (p = .002) and sound-to-picture modality (p < .001), and that the performance in the pictures modality was significantly higher than in the sound-to-picture modality (p = 0.010). In svPPA, the effect of modality on performance was non-significant (*F*(2, 26) = .651, *p* = .530, partial η2 = .048). In sbvFTD, there was a statistically significant effect of modality on performance (*F*(2, 28) = 5.83, *p* = .008, partial η2 = .294). Post-hoc tests showed that performance in the words’ modality was significantly higher than in the pictures’ modality (p = .016), but that all other contrasts were non-significant (Words vs sound-to-picture, p = .263; Pictures vs sound-to-picture, p =.398). In bvFTD, there was a statistically significant effect of modality on performance (*F*(2, 32) = 8.40, *p* = .001, partial η2 = .344). Post-hoc tests showed that performance in the words’ modality was significantly higher than in the pictures modality (p = .023) and sound-to-picture modality (p < .001), but that the performance in the pictures modality did not differ from the sound-to-picture modality (p = 1.000).

#### 3.2.2. Group * Semantic category (Emotions vs Tools)

There was a statistically significant interaction between group and performance depending on the semantic category (*F*(3, 76) = 5.360, *p* = .002, partial η2 = .175) (Figure 1B), controlling for age, sex and education.

In terms of simple main effects of groups, for the emotions’ semantic category, all patient groups performed significantly lower than HC (p < .001 for svPPA and sbvFTD, p=.042 for bvFTD). SvPPA also performed significantly lower than bvFTD (p < .001), but sbvFTD and bvFTD (p = .063) and svPPA and sbvFTD (p = .454) did not significantly differ. For the tools’ semantic category, svPPA and sbvFTD performed significantly lower than HC (p < .001 for both), but bvFTD did not significantly differ from HC (p = 1.000). SvPPA and sbvFTD also performed significantly lower than bvFTD (p < .001 for both).

Finally, svPPA and sbvFTD did not differ significantly (p = .176). Comparisons between patients’ groups were additionally controlled for FTLD CDR Total scores.

In terms of simple main effects of modality, in both HC and bvFTD, there was a statistically significant effect of semantic category on performance, with a significantly better performance in tools knowledge than emotions knowledge (HC: *F*(1, 36) = 41.507, *p* < .001, partial η2 = .536; bvFTD: *F*(1, 16) = 33.93, *p* < .001, partial η2 = .680).

In svPPA, there was no semantic category effect (*F*(1, 13) = .036, *p* =.853, partial η2 = .003), and nor in sbvFTD (*F*(1, 14) = 6.53, *p* < .422, partial η2 = .047).

#### 3.2.3. Group * Condition (Words – tools vs. Words-emotions vs. Pictures – tools vs. Pictures – emotions vs. Sound-to-picture – tools vs. Sound-to-picture - emotions)

There was a statistically significant interaction between group and performance depending on the condition (*F*(15, 380) = 4.934, *p* = <.001, partial η2 = .163) (Figure 1C), controlling for age, sex and education.

For the “Sound-to-picture – Emotions” condition, all patients’ groups performed significantly lower than HC (p < .001 for svPPA and sbvFTD; p =.016 for bvFTD). SvPPA did not differ from sbvFTD (p = .904) and bvFTD (p < .086), and nor did sbvFTD differ from bvFTD (p = .268). For the “Sound-to-picture – Tools” condition, svPPA and sbvFTD performed significantly lower than HC (p < .001 for both), but bvFTD did not significantly differ from HC (p = 367). svPPA performed significantly lower than both sbvFTD (p =.040) and bvFTD (p < .001), and sbvFTD performed significantly lower than bvFTD (p = .041). For the “Pictures – Emotions” condition, all patients’ groups performed significantly lower than HC (p < .001 for svPPA and sbvFTD; p = .006 for bvFTD). SvPPA did not differ from sbvFTD (p = .927). However, both svPPA and sbvFTD differ significantly from bvFTD (p = .040 for svPPA and p = .026 for sbvFTD). For the “Pictures – Tools” condition, svPPA and sbvFTD performed significantly lower than HC (p < .001 for both), but bvFTD did not significantly differ from HC (p = 697). Both svPPA and sbvFTD performed significantly lower than bvFTD (p < .001 for both), but svPPA did not differ from sbvFTD. For the “Words – Emotions” condition, svPPA and sbvFTD performed significantly lower than HC (p < .001 for both), but bvFTD did not significantly differ from HC (p = 854). SvPPA performed significantly lower than both sbvFTD (p = .006) and bvFTD (p < .001). Finally, sbvFTD and bvFTD did not differ significantly (p = .245). For the “Words – Tools” condition, svPPA and sbvFTD performed significantly lower than HC (p < .001 for both), but bvFTD did not significantly differ from HC (p = .923). Both svPPA and sbvFTD performed significantly lower than bvFTD (p < .001 for svPPA and p = .002 for sbvFTD), but svPPA did not differ from sbvFTD. Comparisons between patients’ groups were additionally controlled for FTLD CDR Total scores.

In terms of simple main effects of condition, in HC there was a statistically significant effect of condition on performance, (*F*(5, 180) = 20.949, *p* < .001, partial η2 = .368), with a significantly lower performance in the “Pictures – Emotions” and “Sound-to-picture – Emotions” conditions versus all others (Pictures – Emotions significantly lower than Words – Tools (p=.013), Words – Emotions (p<.001), Pictures – Emotions (p<.001) and Sound-to-picture – Tools (p=.004); Sound-to-picture – Emotions significantly lower than Words – Tools (p<.001), Words – Emotions (p<.001), Pictures – Emotions (p<.001) and Sound-to-picture – Tools (p<.001)). A similar profile was observed in bvFTD: there was a statistically significant effect of condition on performance, (*F*(5, 80) = 10.074, *p* < .001, partial η2 = .544), with a significantly lower performance in the “Pictures – Emotions” and “Sound-to-picture – Emotions” conditions versus all others (Pictures – Emotions significantly lower than Words – Tools (p=.021), Words – Emotions (p=.005), Pictures – Emotions (p<.001) and Sound-to-picture – Tools (p=.009); Sound-to-picture – Emotions significantly lower than Words – Tools (p=.002), Words – Emotions (p<.001), Pictures – Emotions (p<.001) and Sound-to-picture – Tools (p=.002)). In sbvFTD, a significant effect of condition was found (*F*(5, 70) = 6.314, *p* <.001, partial η2 = .311), in which semantic performance was significantly higher in the “Words – Emotions” condition versus all other conditions (p = .044 vs. Words – Tools; p = .023 vs. Pictures – Tools; p = .011 vs Pictures – Emotions; p = .004 Sound-to-picture – Emotions), but the “Sound-to-picture – Tools” condition (p=1.000). Finally, in svPPA, there was no condition effect (*F*(5, 65) = 1.594, *p* =.174, partial η2 = .109).

### 3.3 Brain correlates of the Fear & Spider Test (FST) (Table 3, Figure 4)

Voxel-based morphometry analyses are presented in Table 3 and Figure 4. All results are at a p < .05 FWE cluster-corrected, controlling for age, sex, total intracranial volume and FTLD CDR Total scores, in the full cohort. The two subtasks in the Words modality (Tools, Emotions) showed predominantly left-lateralized patterns, with both subtasks correlating with GM volume in the left ATL and posterior temporal regions, including the inferior, middle, and superior temporal gyri, fusiform gyrus, and parahippocampal cortex. Words–Tools also showed a smaller right ATL cluster, while Words–Emotions extended into the insula and middle frontal gyrus.

**Figure 4.**
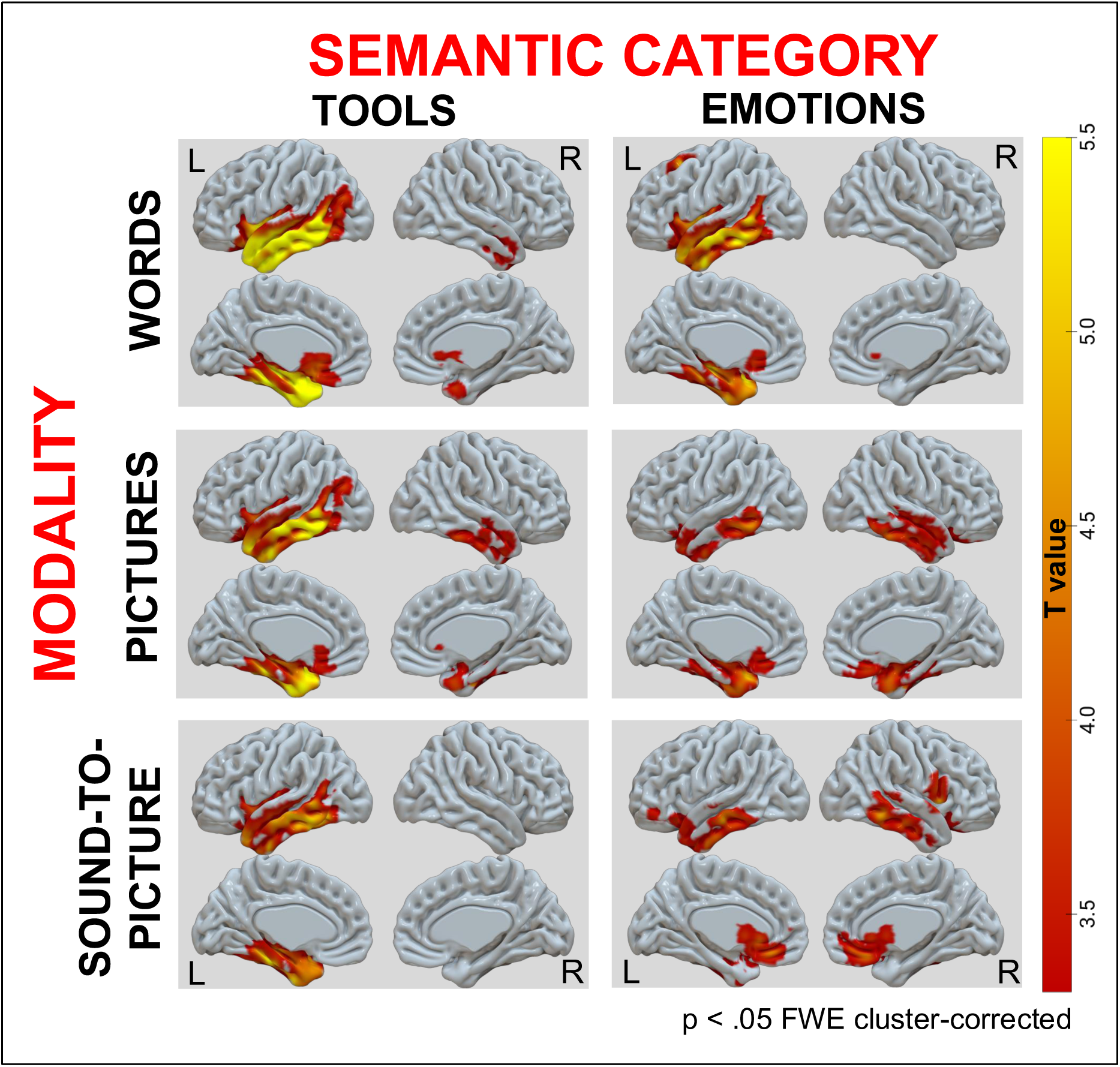
Brain correlates from the Fear & Spider Test (FST) subtasks (p < .05 FWE corrected voxel-level)

**Table 3.** Neuroimaging correlates of the Fear & Spider Test (FST)

| **Region** | **Side** | **x** | **y** | **z** | **Extent** | **T** | **P (corr.) cluster** |
| --- | --- | --- | --- | --- | --- | --- | --- |
| **<u>Words- Tools</u>** |  |  |  |  |  |  |  |
| **Inferior temporal gyrus** | **L** | **-52** | **-4** | **-40** | **44763** | **8.43** | **.000** |
| Middle temporal pole | L | -42 | 10 | -27 |  | 8.33 |  |
| Middle temporal gyrus | L | -64 | -10 | -10 |  | 8.21 |  |
| Inferior temporal gyrus | L | -51 | -48 | -21 |  | 8.07 |  |
| Inferior temporal gyrus | L | -32 | -6 | -44 |  | 7.65 |  |
| Fusiform gyrus | L | -39 | -32 | -24 |  | 7.61 |  |
| Parahippocampal gyrus | L | -28 | -27 | -22 |  | 7.22 |  |
| **Middle temporal pole** | **R** | **40** | **4** | **-32** | **2125** | **4.23** | **.001** |
| Inferior temporal gyrus | R | 46 | -4 | -30 |  | 4.16 |  |
| Middle temporal gyrus | R | 54 | -10 | -21 |  | 3.51 |  |
| Amygdala | R | 24 | 3 | -21 |  | 3.31 |  |
| **<u>Words - Emotions</u>** |  |  |  |  |  |  |  |
| **Hippocampus** | **L** | **-30** | **-22** | **-15** | **31046** | **6.23** | **0.000** |
| Inferior temporal gyrus | L | -52 | -4 | -40 |  | 6.16 |  |
| Inferior temporal gyrus | L | -58 | -38 | -20 |  | 6.13 |  |
| Middle temporal pole | L | -44 | 9 | -26 |  | 5.88 |  |
| Middle temporal gyrus | L | -68 | -10 | -20 |  | 5.81 |  |
| Middle temporal gyrus | L | -66 | -48 | 2 |  | 5.66 |  |
| Superior temporal pole | L | -44 | 2 | -15 |  | 5.53 |  |
| Insula | L | -44 | 10 | -3 |  | 5.52 |  |
| Fusiform gyrus | L | -39 | -38 | -16 |  | 5.27 |  |
| **Middle frontal gyrus** | **L** | **-27** | **18** | **57** | **1181** | **5.19** | **.017** |
| **<u>Pictures Tools</u>** |  |  |  |  |  |  |  |
| **Inferior temporal gyrus** | **L** | **-66** | **-46** | **-15** | **32417** | **7.42** | **.000** |
| Middle temporal gyrus | L | -69 | -34 | -14 |  | 6.79 |  |
| Middle temporal pole | L | -36 | 12 | -32 |  | 6.26 |  |
| Inferior temporal gyrus | L | -32 | -6 | -42 |  | 6.24 |  |
| Superior temporal pole | L | -42 | 8 | -26 |  | 6.15 |  |
| Middle temporal gyrus | L | -62 | -50 | -2 |  | 6.06 |  |
| Fusiform gyrus | L | -27 | -30 | -22 |  | 5.98 |  |
| Fusiform gyrus | L | -20 | 4 | -40 |  | 5.87 |  |
| **Inferior temporal gyrus** | **R** | **48** | **-34** | **-21** | **6627** | **4.5** | **.000** |
| Fusiform gyrus | R | 32 | -12 | -33 |  | 4.39 |  |
| Middle temporal pole | R | 42 | 4 | -32 |  | 4.21 |  |
| Inferior temporal gyrus | R | 46 | -6 | -32 |  | 4.04 |  |
| Superior temporal pole | R | 21 | 9 | -34 |  | 3.94 |  |
| Fusiform gyrus | R | 34 | -26 | -27 |  | 3.92 |  |
| Middle temporal gyrus | R | 57 | -10 | -20 |  | 3.8 |  |
| Parahippocampal gyrus | R | 22 | 2 | -21 |  | 3.59 |  |
| **<u>Pictures-Emotions</u>** |  |  |  |  |  |  |  |
| **Inferior temporal gyrus** | **R** | **51** | **-36** | **-22** | **26525** | **5.09** | **.000** |
| Middle temporal gyrus | L | -39 | 8 | -30 |  | 4.98 |  |
| Inferior temporal gyrus | L | -63 | -50 | -12 |  | 4.81 |  |
| Fusiform gyrus | R | 39 | -38 | -16 |  | 4.69 |  |
| Parahippocampal gyrus | R | 28 | 4 | -34 |  | 4.68 |  |
| Middle temporal gyrus | L | -69 | -34 | -14 |  | 4.57 |  |
| Gyrus rectus | R | 8 | 27 | -22 |  | 4.55 |  |
| Inferior orbitofrontal gyrus | R | 22 | 18 | -22 |  | 4.46 |  |
| Fusiform gyrus | L | -32 | -4 | -39 |  | 4.39 |  |
| **<u>Sound-to-picture - Tools</u>** |  |  |  |  |  |  |  |
| **Inferior temporal gyrus** | **L** | **-51** | **-52** | **-10** | **26604** | **6.41** | **.000** |
| Inferior temporal gyrus | L | -48 | -27 | -26 |  | 6.39 |  |
| Fusiform gyrus | L | -39 | -56 | -12 |  | 6.16 |  |
| Fusiform gyrus | L | -26 | -26 | -26 |  | 6.13 |  |
| Superior temporal gyrus | L | -44 | -6 | -15 |  | 5.38 |  |
| Middle temporal gyrus | L | -64 | -26 | -10 |  | 5.22 |  |
| Inferior temporal gyrus | L | -52 | -4 | -40 |  | 5.21 |  |
| **<u>Sound-to-picture - Emotions</u>** |  |  |  |  |  |  |  |
| **Inferior temporal gyrus** | **L** | **-68** | **-26** | **-18** | **11348** | **5.47** | **.000** |
| Rectus gyrus | L | -3 | 34 | -15 |  | 4.54 |  |
| Insula | R | 24 | 16 | -16 |  | 4.49 |  |
| Inferior temporal gyrus | L | -63 | -50 | -22 |  | 4.32 |  |
| Inferior temporal gyrus | L | -54 | -14 | -34 |  | 4.29 |  |
| Inferior orbitofrontal gyrus | L | -38 | 45 | -14 |  | 4.28 |  |
| Superior orbitofrontal gyrus | L | -24 | 14 | -16 |  | 4.26 |  |
| Superior orbitofrontal gyrus | L | -21 | 40 | -10 |  | 4.21 |  |
| **Superior temporal pole** | **R** | **56** | **10** | **-2** | **1947** | **5.4** | **.003** |
| Rolandic operculum | R | 66 | 3 | 6 |  | 4.54 |  |
| Precentral gyrus | R | 57 | 8 | 20 |  | 4.38 |  |
| **Inferior temporal gyrus** | **R** | **62** | **-45** | **-14** | **4819** | **4.91** | **.000** |
| Middle temporal gyrus | R | 57 | -42 | -4 |  | 4.84 |  |
| Inferior temporal gyrus | R | 57 | -14 | -28 |  | 4.1 |  |
| Superior temporal gyrus | R | 54 | -33 | 9 |  | 3.57 |  |
| Middle temporal gyrus | R | 69 | -21 | -8 |  | 3.42 |  |
| Superior temporal gyrus | R | 64 | -2 | -10 |  | 3.28 |  |

The two Pictures subtasks (Tools, Emotions) showed bilateral patterns of GM correlation. Both tasks involved the bilateral ATL and posterior temporal cortex, but Pictures–Tools showed stronger and more extensive left hemisphere involvement, while Pictures–Emotions was dominated by right-lateralized correlations. These subtasks also involved adjacent regions such as the orbitofrontal cortex and gyrus rectus in the right hemisphere.

For the Sound-to-picture modality, the Emotions subtask was associated with bilateral ATL and posterior temporal GM volume, including the insula and orbitofrontal cortex bilaterally. In contrast, the Tools subtask showed a strongly left-lateralized pattern, correlating exclusively with GM volume in the left ATL and posterior temporal regions, especially within the fusiform and superior temporal gyri.

### 3.4 Subtask-Specific Dissociation in svPPA and sbvFTD (Figure 5; Figure 6)

To test our hypothesis of differences for verbally mediated tool knowledge vs visually mediated emotion knowledge between svPPA and sbvFTD patients, we conducted a focused analysis on two specific conditions: Words–Tools, which we expected to be more impaired in svPPA and associated with left ATL volume, and Pictures–Emotions, which we expected to be more impaired in sbvFTD and associated with right ATL volume.

**Figure 5.**
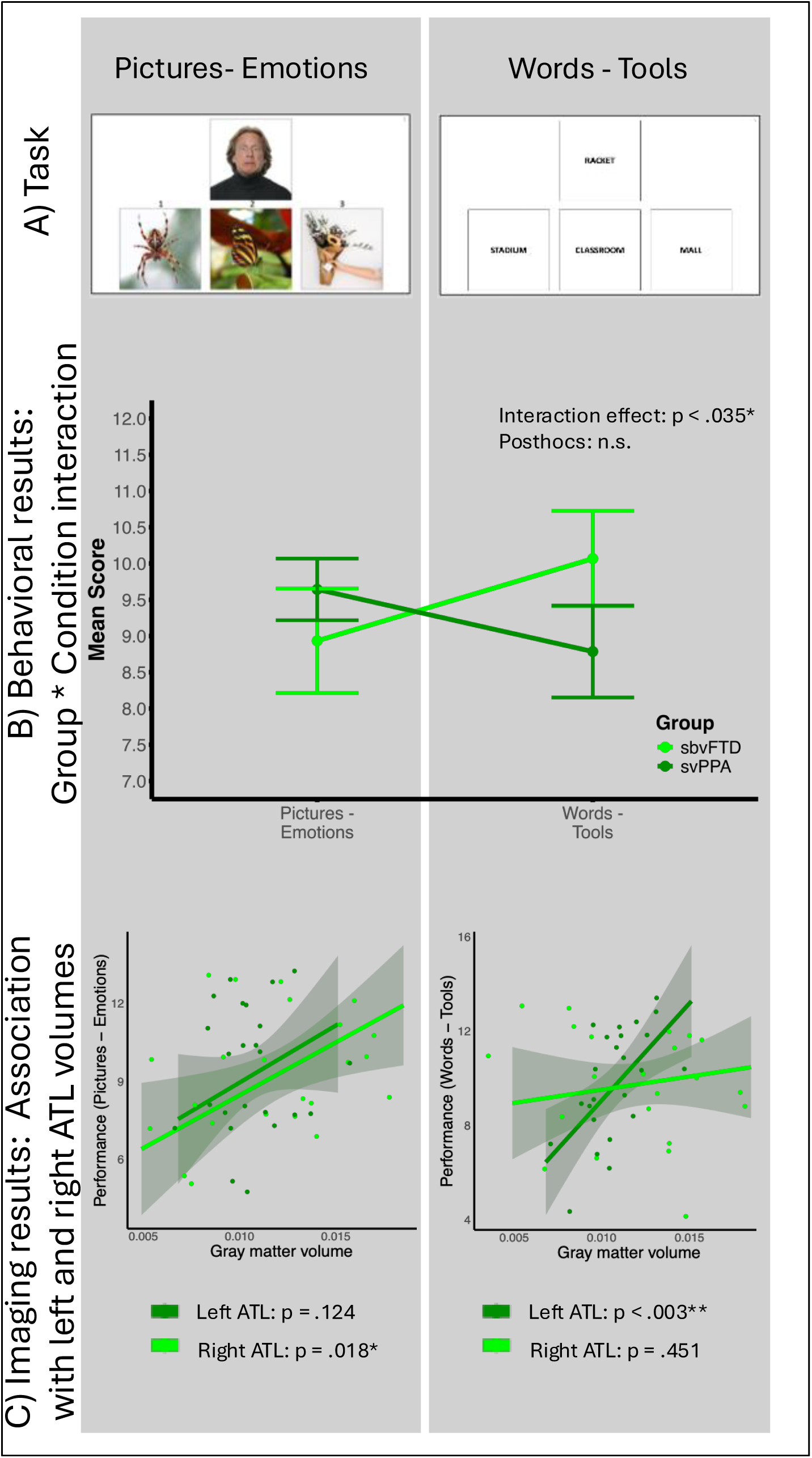
Subtask-Specific Dissociation Between svPPA and sbvFTD: Behavioral Performance and Associations with ATL Volumes

**Figure 6.**
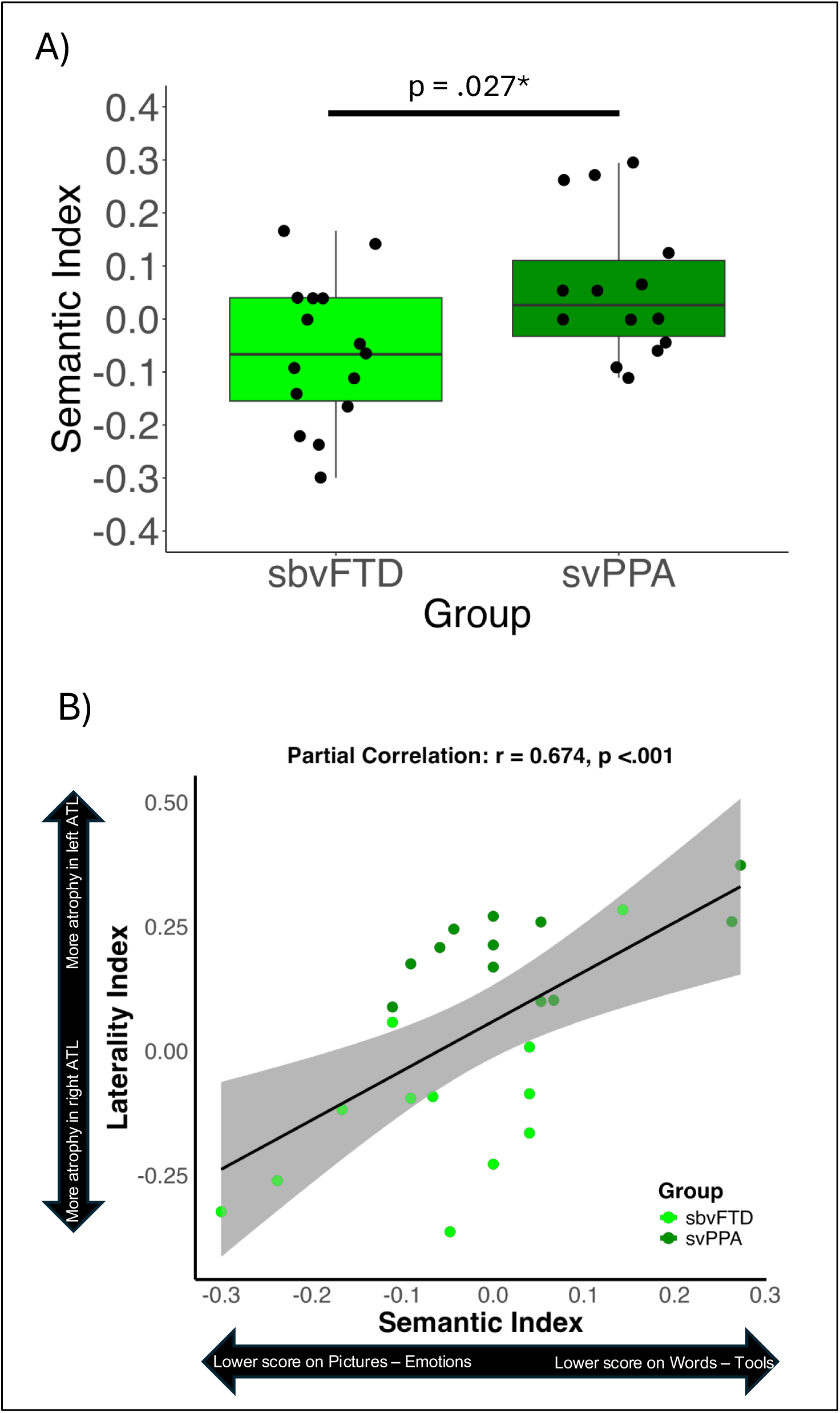
Semantic Index–Based Group Differentiation and correlation with ATL Atrophy Lateralization index

A significant Group * Condition interaction was found (F(1, 26) = 4.945, p < .05, partial η² = .160), controlling for sex (Figure 5A). However, post hoc comparisons within and between groups did not reach statistical significance: svPPA and sbvFTD patients did not significantly differ on either the Words–Tools condition (p = .191) or the Pictures–Emotions condition (p = .393). Similarly, within each group, performance did not significantly differ between the two tasks (svPPA: p = .177; sbvFTD: p = .112).

To move beyond categorical diagnoses, we analyzed the data dimensionally across the full semantic dementia (SD) spectrum. We found that Words–Tools performance was significantly predicted by left ATL volume (p < .01), but not by right ATL volume (p = .451). In contrast, Pictures–Emotions performance was significantly predicted by right ATL volume (p < .05), but not by left ATL volume (p = .124). These models were adjusted for age, sex, and total intracranial volume. Full regression results are provided in Supplementary Tables 1 and 2.

These findings suggest that a composite index integrating both tasks might better capture the different profiles across patients. We thus created a semantic index reflecting relative performance on the two tasks: higher values indicate relatively poorer performance on Words–Tools compared to Pictures–Emotions, while lower values reflect the opposite. Group comparisons revealed that sbvFTD patients had significantly lower semantic index scores than svPPA patients (p < .05), controlling for sex.

Finally, we examined whether this behavioral dissociation aligned with atrophy lateralization. We computed a laterality index reflecting relative atrophy in the left versus right ATL (higher values = greater left ATL atrophy; lower values = greater right ATL atrophy). Across the full SD spectrum, we observed a significant correlation between the semantic index and the laterality index (partial r = .674), controlling for age, sex, and total intracranial volume. The direction of this association indicates that predominant left ATL atrophy is linked to greater impairment in Words–Tools, whereas predominant right ATL atrophy is associated with greater impairment in Pictures–Emotions.

## 4. Discussion

In this study, we examined semantic knowledge using the Fear & Spider Test (FST), a novel multimodal battery contrasting emotional and tool-related concepts across word-based, picture-based, and sound-to-picture modalities in patients with sbvFTD, svPPA, and bvFTD. This test was designed to fill a critical clinical gap by sensitively probing non-verbal emotional semantic deficits in sbvFTD, a recently defined syndrome for which dedicated assessment tools are currently lacking. As expected, patients with both semantic variants showed greater deficits in all semantic tasks compared to controls and bvFTD and bilateral ATL volumes were broadly associated with performance across all semantic tasks. Interactions between modality and categories were necessary for the emergence of differences between sbvFTD and svPPA and right and left ATL atrophy: performance on the Words–Tools condition was more impaired in svPPA and correlated with left ATL volume, while performance on the Pictures–Emotions condition was more impaired in sbvFTD and correlated with right ATL volume. In the following sections, we discuss the clinical, diagnostic implications of these findings and their significance for understanding the neuroanatomical basis of semantic memory.

### sbvFTD and bvFTD: Two Unique Clinical Syndromes Differentiated by Multimodal Semantic Knowledge

Historically, many clinicians and researchers have diagnosed patients with right ATL degeneration as presenting with bvFTD, largely due to the presence of prominent initial behavioral symptoms. This is understandable, given that two of the three core features proposed for sbvFTD, loss of empathy and rigid or compulsive behaviors, are also listed in the bvFTD criteria. This overlap brings us back to the longstanding clinical debate around “lumping” versus “splitting” syndromes. Ultimately, the goal is to predict the underlying neuropathology accurately, and existing evidence shows that sbvFTD and bvFTD are associated with distinct pathological profiles, with sbvFTD being most consistently associated with FTLD-TDP type C (−80%) while bvFTD with more heterogeneous molecular causes (Younes et al., 2022). Therefore, in the absence of FTLD-specific molecular biomarkers, differentiating sbvFTD from the more frontal presentation of classic bvFTD is essential to improve clinical-pathological correlations. Additionally, understanding distinct neural mechanisms underlying behavioral problems could pave the way for future symptomatic therapies, and improved patient management strategies.

Our findings add to this growing body of literature by providing further evidence that these two clinical syndromes are more distinct than previously appreciated. Through a careful evaluation of semantic memory using the Fear & Spider Test, we identified two clearly dissociable cognitive profiles. Nearly all comparisons between the two groups yielded significant differences. Patients with sbvFTD displayed widespread semantic impairments, while those with bvFTD were relatively spared across the board, except in nonverbal subtasks involving emotional concepts. This divergence likely reflects the fact that, when sbvFTD is separated from the broader bvFTD group, the degree of ATL atrophy in bvFTD patients appears less pronounced (see Figure 2). Our results therefore support the utility of assessing multimodal semantic knowledge, particularly of emotions and tools, as a key clinical tool for distinguishing sbvFTD from bvFTD. When combined with other measures such as the recognition and naming of familiar individuals (Yadollahikhales et al., 2024; Younes et al., 2022) and linguistic or prosodic markers from connected speech (Geraudie et al., 2023; Vonk et al., 2025), this approach may offer powerful discriminative capacity for differential diagnosis, as well as improved predictive accuracy regarding the underlying neuropathological processes.

### sbvFTD and svPPA and the Role of the Right and Left ATL in Semantics

Our study shows both overlapping impairments and key dissociations between svPPA and sbvFTD and highlights their relevance to understanding the neural basis of semantic memory. Both patient groups demonstrated widespread semantic impairments. Nonetheless, when considering sensory modality (words, pictures, sound-to-picture) and semantic category (tools, emotions), we found that there was a clear effect of verbal modality when comparing svPPA and sbvFTD. Specifically, sbvFTD patients showed better performance in all verbal subtasks compared to non-verbal ones. This pattern is consistent with the stronger involvement of the right anterior temporal lobe (ATL) in non-verbal semantic processing. Conversely, we did not observe an overall effect of semantic category across all modalities subtasks when comparing sbvFTD and svPPA directly. Instead, significant differences emerged when considering specific interactions based on our specific hypothesis that the left ATL might be more specialized for verbal and tool related features. Consistent with this view, svPPA patients were more impaired in the Words–Tools condition (verbal-tool knowledge), whereas sbvFTD patients showed greater impairment in the Pictures–Emotions condition (non-verbal emotion knowledge). Although direct group comparisons were not statistically significant, likely due to low statistical power, dimensional analyses supported this dissociation and revealed lateralized neuroanatomical correlates: left ATL volume predicted performance in the Words–Tools task, while right ATL volume predicted performance in the Pictures–Emotions task. This suggests that, although svPPA and sbvFTD lie along a shared continuum of semantic impairment in relation to a bilaterally distributed semantic system (M. A. Lambon Ralph et al., 2001, 2010; M. A. L. Lambon Ralph et al., 2017; Rice et al., 2018), carefully crafted tasks that weight on specific semantic features can show differential effects, especially at the initial stages of the disease. To be noted is that, combining scores from these two tasks that do weight on the most likely lateralized combination of category/modality interaction, did allow us to distinguish between svPPA and sbvFTD patients. Testing patients at an earlier stage of the disease, with more extreme levels of atrophy lateralization, may highlight stronger category-specific effects.

In summary, these results reinforce the view that svPPA and sbvFTD represent two clinical syndromes within the broader semantic dementia (SD) spectrum. While each has its own particularities, they share a significant number of symptoms and, most importantly, the strongest prediction of FTLD TDP-43 type C pathological cause. This shared pathology makes clinical differentiation between svPPA and sbvFTD less critical than, for instance, distinguishing sbvFTD from bvFTD. However, studies investigating specific semantic impairments in svPPA and sbvFTD will improve our knowledge of the neural basis of semantic memory in the human brain.

### The Fear & Spider Test: A Novel Clinical Tool

The Fear & Spider Test (FST) offers a flexible and targeted assessment of semantic processing across both sensory modalities and semantic categories. While the test was designed specifically to detect the multimodal semantic deficits characteristic of sbvFTD, and to help differentiate it from bvFTD and svPPA, it may also serve as a useful marker of disease progression in sbvFTD, particularly in longitudinal studies or clinical trials. The FST may prove useful in other clinical contexts as well. For instance, it could be informative in populations where the laterality of ATL atrophy is relevant, such as patients with temporal lobe epilepsy or those who have undergone temporal lobe resection. It may also be valuable for assessing semantic knowledge of emotional cues in clinical populations presenting with social cognition impairments. Although administration can be lengthy in individuals with severe semantic impairments (e.g., semantic dementia), the test is inherently flexible: clinicians can select any subset of the six subtasks depending on the clinical question. Future research should develop a shorter clinical version of the FST, as the current version was designed primarily for experimental use. Nonetheless, like any semantic task, the FST may be influenced by cultural factors, given that emotional concepts can vary across cultural contexts. Further cross-cultural adaptations and normative studies will be necessary to determine whether the tool generalizes to broader populations. Additionally, assessing emotion semantics is inherently challenging, as it is difficult to avoid relying on emotion perception, an ability often impaired in clinical groups. Still, the inclusion of the “Words – Emotions” subtask helps tease apart impairments in emotion reading (e.g., in bvFTD) from broader multimodal emotion knowledge deficits (e.g., in svPPA and sbvFTD). Strengths of the task include the use of facial action coding system (FACS)-validated emotional expressions, the inclusion of both positive and negative emotions, and the careful balancing of stimuli based on culture- and language-specific norms, which were extensively piloted prior to data collection. For example, future trials could examine semantic associations for tools with a greater focus on action-related knowledge (e.g., manipulability) rather than context-related associations, and could include social concepts to help distinguish social knowledge from emotional knowledge.

### 4.7 Limitations

Beyond limitations directly related to the Fear & Spider Test, this study has additional constraints. The relatively small sample size, particularly for sbvFTD, likely limited statistical power and may have obscured some differences between svPPA and sbvFTD. Nonetheless, the dimensional analyses supported this dissociation, indicating that the observed pattern is meaningful even in a modest cohort. Although patients with svPPA and sbvFTD were at relatively early stages of disease, the presence of bilateral anterior temporal lobe atrophy may have reduced sensitivity to subtle differences in semantic performance. Earlier identification of these syndromes could facilitate future studies focused on even milder clinical presentations. Although we statistically controlled for demographic variables and disease severity, unequal sex distribution across groups may have introduced residual confounding. Furthermore, volume-based correlations are more likely to detect brain–behavior relationships in regions that are affected by atrophy in the sample and may be less sensitive in relatively spared regions. Finally, the sample was composed primarily of white, highly educated, native English speakers. Broader sampling in future studies will be critical to determine whether these findings generalize across different cultural, linguistic, and socioeconomic backgrounds.

### 4.8 Conclusion

Our findings demonstrate that FTD syndromes show differences in semantic impairments with strong effects based of modality of input and more nuanced effects of semantic category. This study further underscores the distinctiveness of sbvFTD as a clinical syndrome, one that is meaningfully different from bvFTD and aligns more closely with svPPA under the broader SD umbrella. As sbvFTD becomes increasingly recognized, the Fear & Spider Test provides a valuable addition to the growing arsenal of tools available to detect and characterize this variant in clinical settings. By strengthening the clinical distinction between syndromes with distinct underlying pathologies, our work lays essential groundwork for the development of more precise, pathology-targeted diagnostic and therapeutic strategies.

## Data Availability

While we can share anonymized data, public archiving is not yet permitted under the study’s institutional review board approval due to the sensitive nature of patient data. Specific requests can be submitted through the UCSF – MAC Resource (Request form: http://memory.ucsf.edu/resources/data). Following a UCSF-regulated procedure, access will be granted to designated individuals in line with ethical guidelines on the reuse of sensitive data. This would require submission of a Material Transfer Agreement, available at: https://icd.ucsf.edu/material-transfer-and-data-agreements. Commercial use will not be approved.

---

**Supplementary figure 1.**
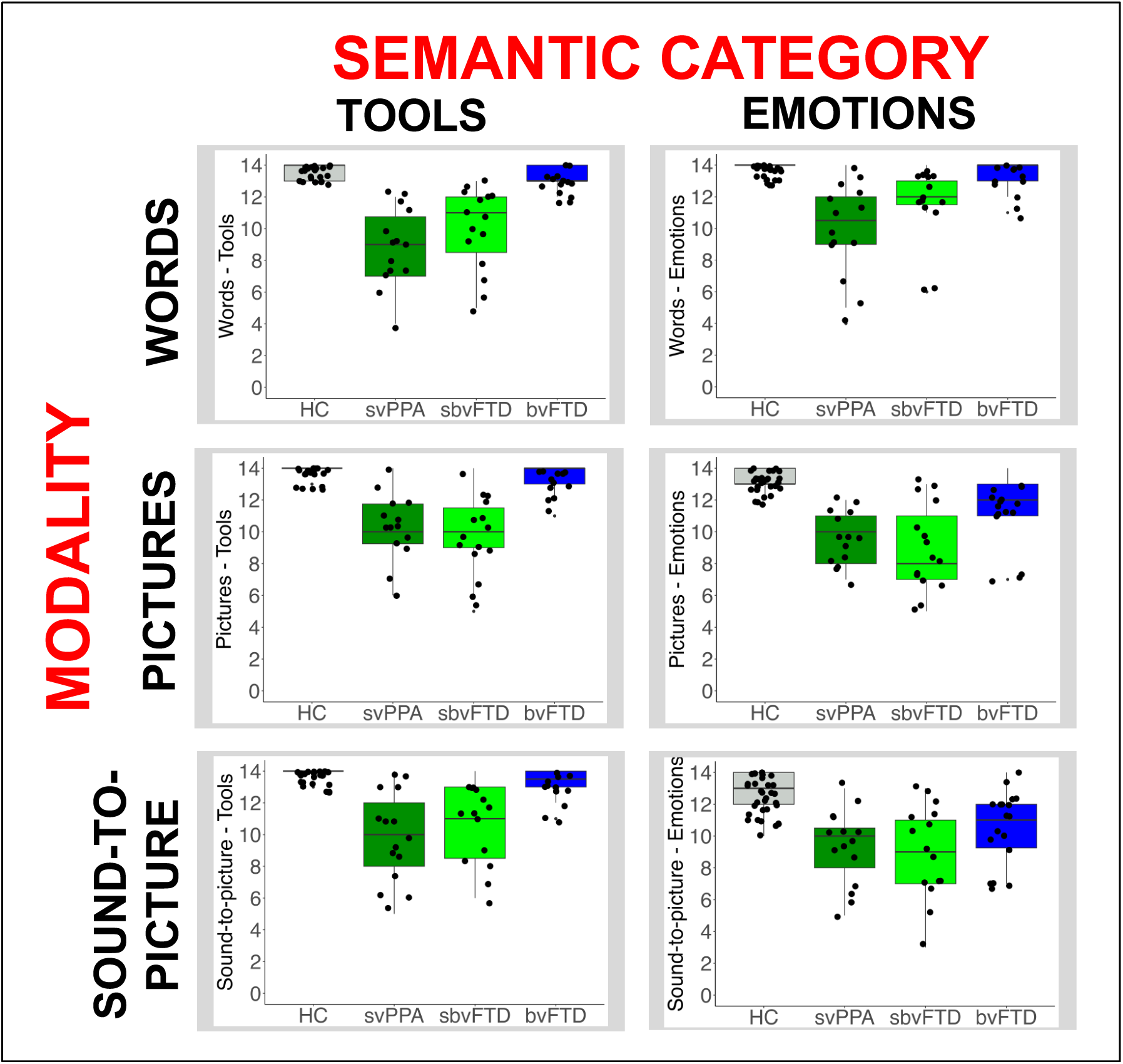
Boxplots of the behavioral results from the Fear & Spider test (FST).

**Supplementary table 1.**
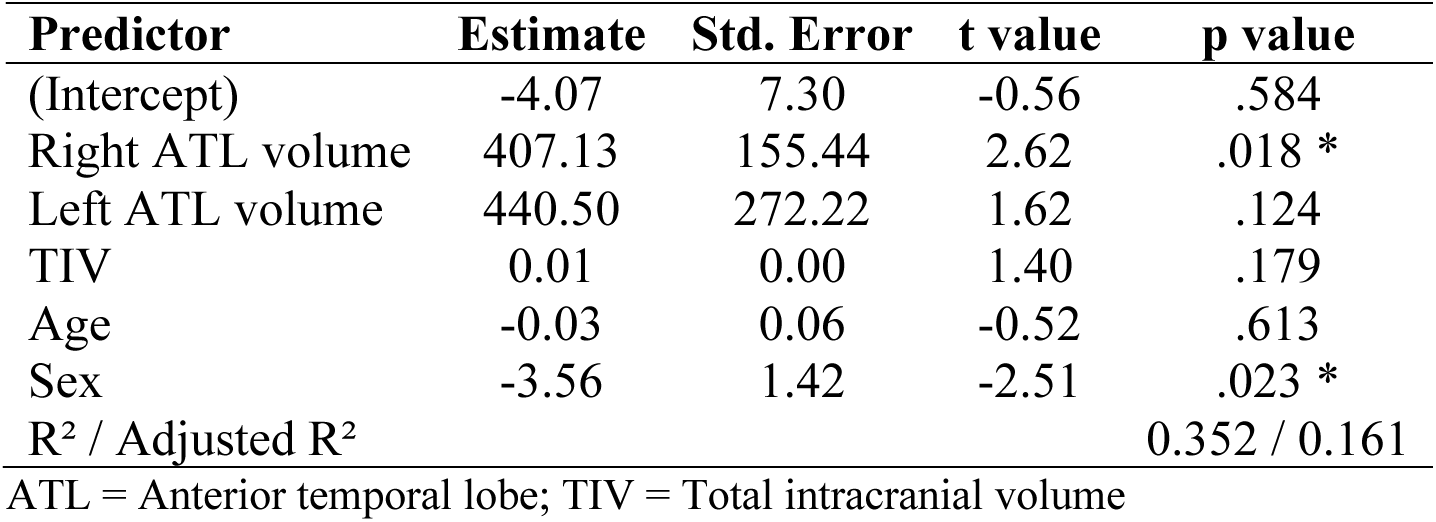
Regression model predicting performance on Pictures-Emotions in SD patients only (svPPA & sbvFTD)

**Supplementary table 2.**
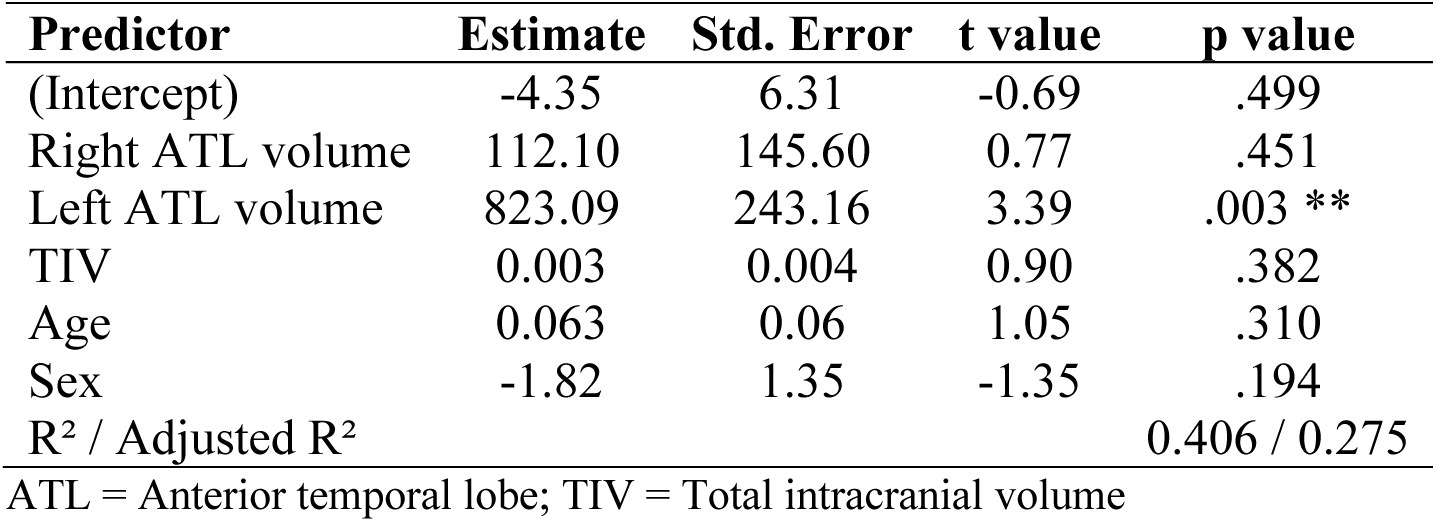
Regression model predicting performance on Words - Tools in SD patients only (svPPA & sbvFTD)

